# An EBV-associated atypical B cell signature in clinically isolated syndrome is implicated in progression of multiple sclerosis

**DOI:** 10.1101/2023.02.26.23286433

**Authors:** Elliott D. SoRelle, Ellora Haukenfrers, Vaibhav Jain, Karen Abramson, Emily Hocke, Laura A. Cooney, Kristina M. Harris, Scott S. Zamvil, Simon G. Gregory, Micah A. Luftig

## Abstract

Expansion and pathogenicity of CD19^+^/CD20^+^/CD11c^+^/T-bet^+^ atypical B cells (ABCs) are hallmarks of numerous autoimmune disorders and chronic infections. In many such cases Epstein-Barr virus (EBV) is another associated or etiologic factor, though EBV involvement in these diseases remains poorly understood. Notably, the expansion of pro-inflammatory ABCs and a putative causal role for EBV have been identified independently in multiple sclerosis (MS). A common precipitating event in MS onset is Clinically Isolated Syndrome (CIS), a neuroinflammatory demyelinating condition of which 60-80% of cases progress to relapsing-remitting MS (RRMS). Here we report single-cell gene and surface protein expression (scRNA/CITE-seq) in peripheral B cells collected longitudinally from patients with CIS during the Immune Tolerance Network STAyCIS Trial. We focus on the transcriptomic signatures of ABCs from this cohort, publicly available scRNA-seq datasets from six other autoimmune and chronic infectious diseases, and *in vitro* EBV infection. Conservation of an expanded ABC expression profile across diseases establishes ABC dysregulation as a feature of CIS. Critically, we also observed transcriptomic features that distinguished CIS and *de novo* EBV-infected ABCs from those found in healthy controls and other disease contexts. Outcome stratification of CIS samples revealed a rare yet distinctive pro-inflammatory ABC subset that was significantly underrepresented in long-term non-progressor (LTNP) versus cases with RRMS activity (∼5-fold difference). Collectively, this study provides evidence for altered ABC regulation – possibly arising from niche-specific responses to EBV infection – preceding MS onset.

**SUMMARY:** Single-cell transcriptomics establishes an EBV-associated signature in T-bet^+^ atypical B cells in CIS and a pro-inflammatory phenotype underrepresented in patients with no disease progression.

## MAIN

Atypical B cells (ABCs) represent a unique immune compartment broadly identified by co-expression of the canonical T cell lineage transcription factor T-bet (encoded by *TBX21*), the plasmacytoid Dendritic Cell marker CD11c (*ITGAX*), and the classical B cell markers CD19 and CD20 (*MS4A1*)^1, 2^. Originally discovered in mice as an age-associated immune subset, these cells accumulate in response to toll-like receptor 7 (TLR7) and interferon gamma (IFNγ) stimulation and exhibit B cell receptor (BCR) anergy in addition to molecular indicators of exhaustion, inhibited adaptive response, and extrafollicular development^1,3–8^. ABC pools in mice and humans contain high frequencies of autoreactive clones and are expanded in genetic females due at least partially to dosage effects of TLR7, which is encoded on the X chromosome in each species and escapes X-linked inactivation^1,5,9–11^. Although they are present in healthy individuals and can mediate protective immune responses^12–14^, T-bet^+^ B cells co-expressing combinations of other markers (e.g., FCRL5, CXCR3) are common pathogenic effectors in autoimmune and infectious diseases, many of which exhibit a sex bias toward females^15,16^. These include systemic lupus erythematosus (SLE)^9,17–19^, primary Sjögren’s Syndrome (PSS)^20^, rheumatoid arthritis (RA)^21,22^, and chronic infections including human immunodeficiency virus (HIV) and malaria^23–28^.

Epstein-Barr virus (EBV) is a gammaherpesvirus that establishes lifelong infection in more than 90% of adults worldwide^29^. This striking prevalence is largely attributable to the success with which EBV induces and modulates the adaptive immune responses of host B cells to achieve persistent latency in the memory B cell pool via germinal center (GC) dynamics as well as GC-independent routes^30–33^. In addition to its contribution to the development of multiple cancers^34^, EBV is frequently associated with each of the autoimmune and chronic infectious diseases that exhibit clonally expanded ABCs^35–46^. We recently reported that EBV can infect existing ABCs *de novo*^33^ and promote their formation in models of latent infection *in vitro*^47^. However, the manifestation and mechanistic contributions of EBV infection in autoimmune and chronic infectious diseases – including whether they are cell niche-dependent – are incompletely understood.

Multiple sclerosis (MS) is a chronic neuroinflammatory disease with a 3:1 female bias^48^ in which both pathogenic ABCs^49,50^ and epidemiologic evidence of EBV etiology in disease onset (a 32-fold risk from EBV seropositivity after childhood) have been identified^51,52^. Several mutually inclusive hypotheses have been proposed to explain EBV involvement in MS. These include immune responses targeted toward viral proteins that are cross-reactive with self-antigens (molecular mimicry), EBV-induced licensing of (forbidden) autoreactive B cell clones, and damage to bystander cells caused by pro-inflammatory responses of infected cells^53–55^. For example, antibody reactivity and T cell specificity profiles from patients with MS provide evidence for molecular mimicry between the viral latency protein EBNA1 and several CNS-expressed host proteins^56,57^. CNS inflammation originating from myeloid cell activation by a distinct GM-CSF^+^ B cell subset has also been described^58^. Moreover, multiple EBV antigens and EBV^+^ B cells have been detected in MS brain lesions, though not consistently across studied cases^59–64^. Intriguingly, a correlation between neuroinvasive CXCR3^+^ B cells and EBV viral load has been identified recently in patients with MS.^65^ In addition to shedding light on the clinical efficacy of B cell depletion therapy, these and related findings to date underscore substantial consequences of immune dysregulation mediated by EBV in B cells within the CNS. Exactly how the virus gains access to the CNS and the extent to which EBV promotes pathogenesis via the mechanisms described above are open questions. However, these findings and numerous clinical aspects of MS are consistent with the molecular markers and functions of ABCs and suggestive of a role for EBV infection within this niche.

Clinically isolated syndrome (CIS) is an initial clinical episode of neurologic symptoms caused by neuroinflammation and demyelination^66^ that commonly precedes MS. Patients with CIS that also present with brain or spinal cord lesions detected by magnetic resonance imaging (MRI) have a 60-80% chance of subsequent neurologic events and diagnosis with RRMS^67–69^. Despite its diagnostic timeline and significant clinical relation to MS, the presence, frequency, and phenotypic characteristics of ABCs in CIS cohorts have not been studied, to our knowledge. Similarly, whether signatures of ABC response to EBV infection are present in CIS or across the spectrum of virus-associated diseases described above^33^ is not known. Thus, we aimed to transcriptomically profile ABCs from patients with CIS and compare them with ABC compartments from other disease cohorts as well as data from *in vitro* EBV infection.

Peripheral blood mononuclear cells (PBMCs) were collected from a cohort of patients (n = 16; 11 female, 5 male) at two timepoints following initial CIS diagnosis: t_1_ (baseline visit; at least 28 days after completion of corticosteroid treatment and within 210 days of CIS presentation) and t_2_ (at least 3 months post-t_1_; mean = 172 days post-t_1_, range = 84-357 days). Beginning at t_1_, eleven patients were treated with atorvastatin and five were given a placebo as part of the Immune Tolerance Network (ITN) STAyCIS trial^70^ (NCT00094172). Treatments were given for 12 months or until a participant met the primary endpoint of MS activity, defined as 3 or more new T2 lesions or one clinical exacerbation. Eight of the 16 participants met the primary endpoint within the 12 month treatment window, and two additional participants met the primary endpoint in a follow-up period between 12 and 18 months. Of the remaining six participants, three were subsequently diagnosed with MS on the basis of new T2 lesions. Thus, 13 of 16 participants in the analyzed cohort were diagnosed with MS. The remaining three participants were characterized *post hoc* as long-term non-progressors (LTNP) based on the collective absence of new T2 lesions, Gd-enhancing lesions, and clinical exacerbations in the 18 months following t_1_. Of 13 individuals diagnosed with MS, ten received atorvastatin and three received placebo; of three LTNP individuals, two received atorvastatin and one received placebo. PBMCs from each patient and timepoint were prepared as single-cell libraries (scRNA-seq + CITE-seq), which were sequenced, quality controlled, and aligned to generate count matrices. Single-cell data from patients with CIS were then analyzed in conjunction with publicly available scRNA-seq datasets from autoimmune (MS, SLE, PSS, RA)^49,71–74^ and chronic infectious diseases (HIV, malaria)^26^ in which pathogenic ABCs have been described (**Fig. 1A**). Peripheral B cells were identified from each dataset based on *CD19* and *MS4A1* (*CD20*) co-expression and isolated for downstream analysis. We also incorporated data from ABCs prior and subsequent to EBV infection *in vitro*^33^ to assess transcriptomic similarities among disease and EBV^+^ ABCs (**Fig. 1B**). *CD19^+^*/*MS4A1^+^* expressing clusters from our CIS and publicly available datasets were merged into an object containing over 60,000 cells from seven diseases, healthy patient controls, and pre-vs-post EBV infection *in vitro* (**Fig. 1C**). The CIS data reported herein contain 20,839 annotated B cells collected from individuals across the two timepoints (n = 32). Of these, we focused on a population of 5,926 cells found via low-resolution unbiased clustering to contain cells expressing the ABC marker *TBX21* (T-bet). While the number of patients from publicly available datasets varied (n_HIV_ = 3; n_Malaria_ = 3; n_RA_ = 4; n_PSS_ = 5; n_MS_CSF_ = 24, n_MS_PBMC_ = 5; n_SLE_adult_ = 7; n_SLE_child_ = 33), a minimum of ∼2,000 B cells per disease and healthy controls (n_adult_ = 8; n_child_ = 11) were analyzed. Next, unsupervised methods were used to define 42 clusters for preliminary analysis (**Fig. 1D**). At this resolution, elevated expression of *TBX21* and *CXCR3* (another marker of pathogenic ABCs) was observed in approximately six of eleven SLE, four of five CIS, six of seven MS, two of three HIV, two of four PSS, one of six RA, two of four malaria, and *in vitro* EBV^+^ clusters (**Fig. 1E**).

**Figure 1.**
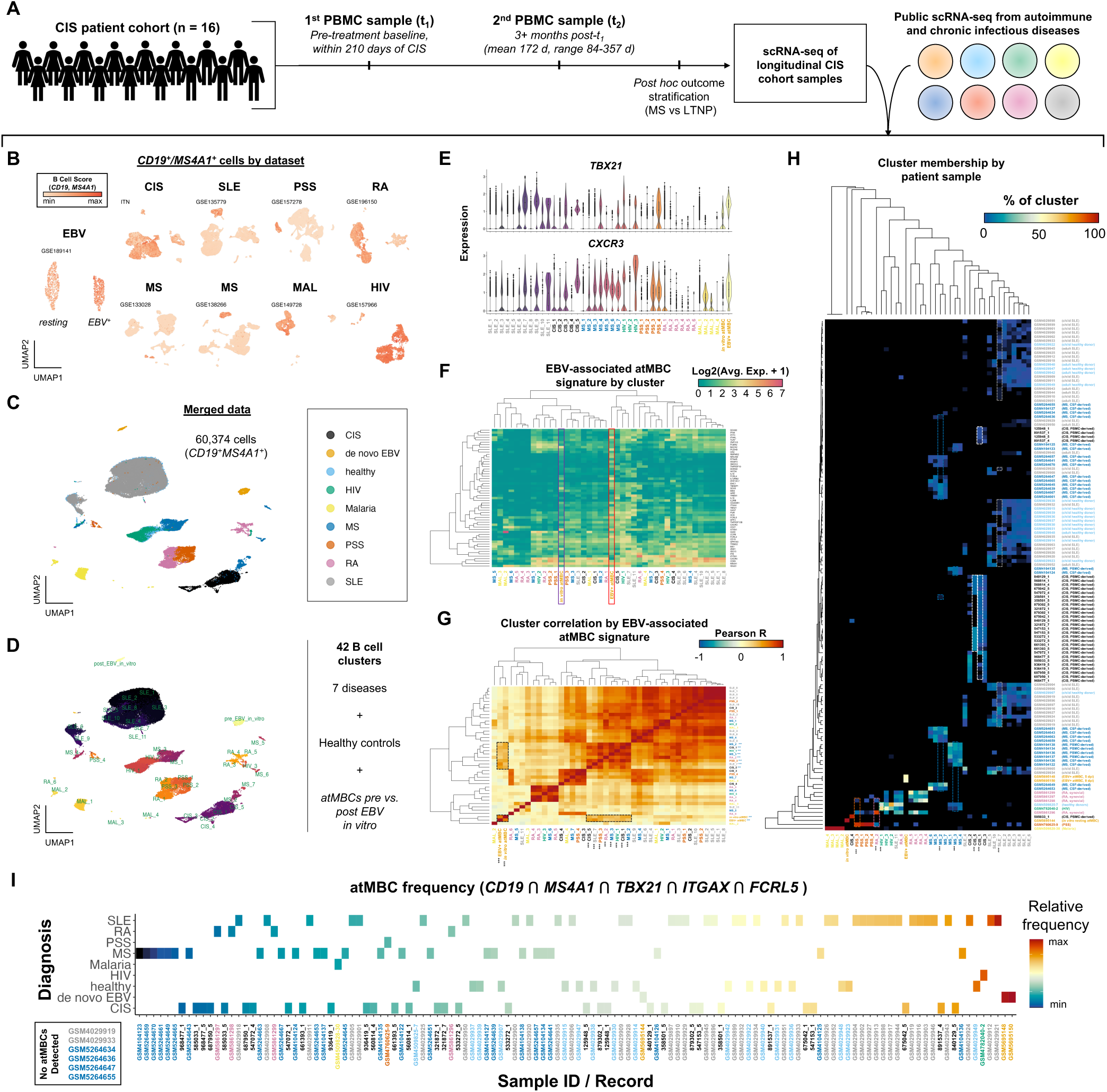
Survey of ABCs in autoimmune and chronic infectious disease scRNA-seq datasets. **A)** Study overview: single-cell sequencing data were collected from a longitudinal CIS cohort (ITN STAyCIS Trial) and jointly analyzed with publicly available scRNA-seq datasets from autoimmune and chronic infectious diseases. **B)** Scoring of classic B cell lineage gene expression (*CD19*, *MS4A1* (CD20)) by single-cell dataset. **C)** UMAP of merged B cells identified across single-cell datasets annotated by disease. **D)** UMAP of merged B cells annotated by clusters determined through unsupervised methods. Cluster names reflect original disease context. **E)** Expression of *TBX21* (T-bet) and *CXCR3* in B cells by disease clusters depicted in (D). **F)** Hierarchical ordering of B cell clusters by average expression of gene signature associated with EBV infection of ABCs from *in vitro* studies. Expression from ABCs before and after *de novo* EBV infection *in vitro* are highlighted with black and red boxes, respectively. Cluster dendrograms depict expression similarity across genes (rows) and similarity across disease clusters (columns). **G)** Pearson correlation matrix of disease B cell clusters by expression of EBV-induced gene signature. Dashed boxes and triple asterisks (***) denote ABC clusters from *de novo* infection study (EBV^+^ and EBV^-^) and B cell clusters from disease datasets with the strongest correlation to these phenotypes. **H)** Patient sample composition of disease B cell clusters. Dashed boxes highlight patient sample representation in select clusters with high correlation to EBV^+^ ABCs. Heatmap color encodes the percentage of cells within each cluster derived from patient-level sequence records. CIS samples are listed by de-identified patient records defined in this study; samples from publicly available datasets are listed by GEO submission ID. **I)** Frequency of ABCs by disease and patient record determined by co-expression of *CD19*, *MS4A1*, *TBX21* (T-bet), *ITGAX* (CD11c), and *FCRL5*.

We next performed hierarchical ordering of the 42 *CD19*^+^/*MS4A1*^+^ clusters based on average expression of genes of interest previously identified in EBV^+^ atMBCs (**Fig. 1F**). These included ABC markers (*TBX21, CXCR3*, *FCRL5*, *SOX5*); interferon-induced and antiviral responses genes (e.g., *DDX60*, *IFI6*, *IFI44*, *IFI44L*, *IFIT3*, *ISG15*, *MX1*, *TBKBP1*, *TRIM22*); early markers of B cell or other immune subset activation (e.g. *CD69*, *CCR6*, *GRP183*, *TNFRSF19*, *TNFRSF13B*); cytokines and other inflammatory response mediators (e.g., *IL18*, *CD200R1*, *FGR*, *HCK*); and a number of EBV-induced genes associated with neuronal gene ontology (e.g., *ENC1*, *NRCAM*, *PPP1R17*, *RTN4R*)^33^. Notably, *in vitro* EBV^+^ ABCs clustered more closely to several disease cell phenotypes than cells prior to infection. Pairwise cluster correlation based on these markers was not strongly disease-dependent, confirming similarities between EBV-infected cells and those from CIS_5, CIS_1, SLE_7, PSS_3, RA_2, MS_2, MS_3, and HIV_1 (**Fig. 1G**). While a limited number of samples were analyzed for Malaria, PSS, RA, and HIV, patient representation within each cluster clearly showed that CIS, MS, and SLE cell similarities with EBV^+^ ABCs were not artificially driven by only one or a few individuals (**Fig. 1H**). Nearly 90% of CIS patient datasets contained cells matching the CIS_1 phenotype (28 of 32 samples), whereas a much smaller cluster, CIS_5, was derived from 11 of 32 patient samples. Roughly 30-50% of 29 analyzed MS patient datasets included cells found within the MS_1 and MS_2 clusters (9 of 29 and 15 of 29 records, respectively). The SLE_7 cluster contained cells from 75% (30 of 40) analyzed datasets from adults and children with SLE. Enumeration of cells co-expressing the ABC markers *CD19*, *MS4A1*, *TBX21*, *ITGAX* (CD11c), and *FCRL5* by sample highlighted the variable frequency of ABCs in disease (**Fig. 1I**). The highest frequencies of ABCs were measured in patient samples from SLE and HIV infection. ABCs were more frequent in patients with CIS than in those with MS, although this may be partially due to the majority of analyzed MS samples being derived from CSF rather than PBMCs.

We aimed to establish deeper phenotypic profiles of ABCs identified in each disease. To do so, we extracted disease clusters with similarity to EBV^+^ ABCs (CIS_5, CIS_1, SLE_7, PSS_3, RA_2, MS_2, MS_3, and HIV_1) and refined them through additional clustering (**Figs. 2A, S1**). This yielded at least one *TBX21^+^*ABC subset per disease and *in vitro* EBV infection. Single-cell co-expression of *TBX21*, *CD19*, *MS4A1*, *FCRL5*, *ITGAX*, and *SOX5* confirmed hallmark ABCs in each disease, including multiple clusters from patients with CIS (**Fig. 2B**, left top panel). Expression of the transcription factor *FOXP4* was also conserved in ABCs. Notably, ABCs exhibited variable *CXCR3* expression across diseases. This variation may reflect origins from distinct B cell lineages^75^ or resting (*CXCR3*^lo^) versus stimulation-induced activated or differentiated (*CXCR3*^hi^) phenotypes^47,76,77^. We also confirmed expression of top differentially expressed markers of ABCs previously identified in publicly available datasets (*ITGB2*, *MPP6*, *NEAT1*, *PLEK*, *S100A11*, and *TNFRSF1B*) within CIS cohort and *de novo* EBV infection samples (**Fig. 2B**, left middle panel).

**Figure 2.**
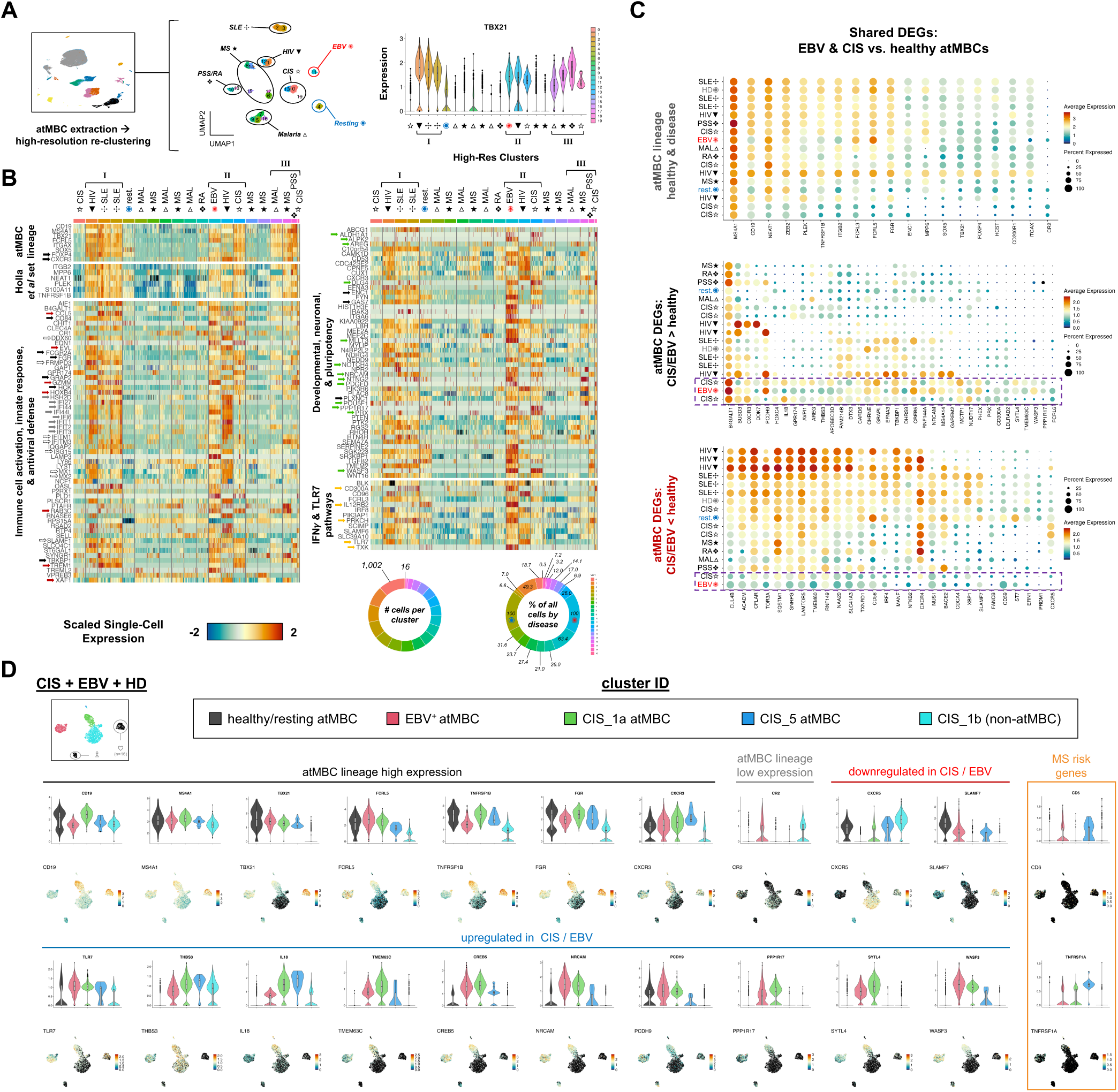
Comparison of ABC transcriptomes by disease identifies shared signature in CIS and *de novo* EBV infection. **A)** Extraction and high-resolution re-clustering of B cells with ABC phenotypes across disease contexts (left and middle panels). *TBX21* expression after re-clustering to identify strongest ABC signatures annotated by disease (right panel). **B)** Single-cell gene expression in high resolution clusters. Heatmap panels present expression of lineage hallmarks and top markers of ABCs identified in previous studies as well as EBV-induced differentially expressed genes involved in immune cell activation (black arrows), innate (gray arrows) and inflammatory responses (red arrows), antiviral defense (dashed arrows), neuronal ontology (green arrows) and developmental pluripotency, and interferon gamma and TLR7 pathways (gold arrows). **C)** Expression of genes with shared expression trends in ABCs in CIS and *de novo* EBV infection contexts relative to healthy donors across high-resolution disease clusters. Select ABC lineage genes (top panel), genes with elevated expression in CIS and *de novo* EBV contexts (middle panel), and genes with reduced expression in CIS and *de novo* EBV contexts (bottom panel) are presented by expression level and frequency of positive cells. **D)** Detailed expression of select genes with common trends in CIS and *de novo* EBV-infected ABC subsets versus healthy donor ABCs and *TBX21*^-^ B cells from CIS samples. Violin and UMAP expression plots for ABC lineage genes (black & gray brackets), upregulated genes (blue bracket), downregulated genes (red bracket), and genes associated with MS risk.

We also assayed co-expression of markers from significantly enriched gene ontology (GO) sets, with particular focus on those upregulated upon *de novo* EBV infection of ABCs^33^ (**Fig. 2B**). ABCs in each disease displayed higher expression of genes involved in immune cell activation, innate response, and antiviral defense (**Fig. 2B**, left bottom panel). For example, *CD84*, *FCGR2A* (CD32), the EBV EBNA-2 target *FGR*^78,79^, *GRAP2*, *HCK*, and *TBKBP1* were consistently and specifically expressed by ABCs in every examined context. Others were enriched in the niche across most diseases (*MX1*, *MX2; FRMPD3*, *IFITM1*, and *IFITM3,* except in Malaria; *DDX60* and *SLAMF1* (CD150), except in MS; *ISG15,* except in PSS and RA). Genes that were generally expressed in, but not specific to, ABCs included *HSH2D*, *IFI27*, *IFI44*, *IFI44L*, and *IFI6* (except in MS, PSS, and RA); *IFIT1,* and *IFIT3* (except in MS, Malaria, PSS, and RA); and *IFIT2* (except in MS). Expression of certain genes was largely restricted to ABC clusters in a subset of diseases. These include *CCL5*, *EVL*, *GZMM*, *HOXB4*, *RAB3C*, *TREM1*, and *XAF1*. Finally, a limited number of genes with functions in innate immune response displayed comparatively disease-specific expression in ABCs (*AIF1* (IBA1), *GPR174*, and *OASL* in SLE and CIS; *LYST*, *RSAD2* in HIV and SLE; *PTAFR* in HIV and MS; and *RTP4* in HIV and CIS).

Genes with annotated developmental, cell plasticity, or neuronal lineage ontologies that we previously identified in response to EBV infection were expressed by ABCs in one or more autoimmune and chronic infectious diseases (**Fig. 2B**, right top panel). Most of these genes are infrequently expressed in mature B cells and their products support pluripotency (*ALDH1A1*, *POU5F1*, *MLLT3*, *NOTCH4*); cell motility and migration (*CXCR3, WASF3*); inflammatory and pro-fibrotic responses (*AREG*, *NOTCH4*, *PDGFD*); and neural cell specification and function (*ENC1*, *GAS7*, *NRCAM*, *NTNG2*, *PLXNC1*, *PPP1R17*, *PRX*). Elevated expression of several of these genes (*ENC1* and *GAS7*) has been identified previously in ABCs, particularly those from patients with SLE^19,76^. Based on their functional importance to ABC responses, we also examined genes with annotated roles in IFNγ and TLR7 signaling (**Fig. 2B**, right bottom panel). Within these GO sets, *FCRL3*, *PIK3AP1*, *SCIMP*, and *SLAMF6* were upregulated across ABCs in disease. *TLR7* expression, which is known to be induced upon EBV virion entry into host B cells^80^, was upregulated in ABCs from HIV, SLE, and CIS. Distinctive expression in ABC subsets was observed for *CD300A* (SLE, CIS); *PRKCH* (SLE, CIS, MAL, PSS, and RA); *IL12RB2* (SLE, CIS); and *TXK* (CIS). *IRF8* was enriched within ABCs in HIV and to a lesser degree in MS and SLE. Collectively, this analysis reinforces prior hypotheses and findings that pathogenicity of ABCs in autoimmunity and chronic infection may be mediated by common mechanisms^26,81^. Moreover, the presence and extent of shared gene expression trends between disease-state and EBV-infected ABCs suggest a potential role for EBV in mediating or inducing ABC pathogenesis.

Curiously, the expression of certain genes (e.g., *ALDH1A1*, *ALPK2*, *DLG4*, *MLLT3*, *PDGFD*, *PPP1R17*, *PRX*, *WASF3*) was virtually exclusive to CIS and *de novo* EBV-infected ABCs. Given this observation, we further investigated genes with shared expression trends in CIS and *de novo* EBV-infected versus healthy donor ABCs in other diseases (**Figs. 2C-D, S2**). Expression of established biomarkers (*CD19^+^*/*MS4A1^+^*/*TBX21^+^*/*ITGAX^+^*/*FCRL5^+^*/*SOX5^+^*/*CR2^-^*) and other conserved transcriptomic features of ABCs (*TNFRSF1B*, *ITGB2*, *FCRL3*, *CD200R1, ZEB2*, *FOXP4, ENC1*, *FGR*, *HCST, MPP6*, *PLEK, NEAT1*) provided a reference for differential expression trends specific to CIS (open stars) and EBV^+^ (red asterisk) ABCs (**Fig. 2C**, top panel). Genes that were commonly enriched in CIS EBV-infected ABCs relative to healthy controls (gray and blue dotted circles) included *SUSD3* and *CXCR3* (also up in MS, PSS, HIV, and SLE); *PCDH9* (also up in PSS and HIV); *AREG* (up in malaria and RA); *CREB5* and *NRCAM* (also up in SLE, HIV, and malaria); *MS4A14* and *GAREM2* (also up in HIV); and *AVPI1*, *CD300A*, *FCRL6*, *GPR174*, *HOXC4*, *IL18*, *NUDT17*, *PPP1R17*, *SYTL4*, *THBS3*, *TMEM63C*, and *WASF3* (up in CIS and *de novo* EBV only) (**Fig. 2C**, middle panel). Conversely, genes that were downregulated in CIS and EBV-infected ABCs included *IRF4* and *MANF* (also down in MS, malaria, PSS, and RA) as well as *BACE2*, *CDCA4*, *CXCR5*, *NUS1*, *SLAMF7*, *ST7*, and *XBP1* (also down in ABC subsets from HIV, malaria, MS, and RA) (**Fig. 2C**, bottom panel).

To further focus our comparison of CIS and EBV transcriptomic signatures, we merged ABC clusters from healthy donors (HD, n = 16), pre-EBV infection *in vitro* (resting), post-EBV *in vitro*, two CIS atypical states (CIS_1a, CIS_5) and one CIS *TBX21^-^* state (CIS_1b, representing 15 of 16 CIS patients) (**Fig. 2D**). Beyond the conservation of lineage-defining markers in healthy, EBV-infected, and CIS ABCs, reduced expression of *SLAMF7* (*CD319*, *CRACC*) occurred in response to EBV infection and in CIS (in both atypical and conventional B cells) relative to cells from healthy donors. While *SLAMF7* expression upon conventional B cell activation was demonstrated to promote proliferation and cytokine expression^82^, it negatively regulates pro-inflammatory responses in NK cells^83^. Reduced *CXCR5* and variable *CXCR4* expression in EBV^+^ and CIS ABCs was also consistent with prior studies^23,84–86^. Notable genes with higher expression in EBV^+^ and CIS patient ABCs relative to healthy controls included the pro-inflammatory (IFNγ-inducing) cytokine *IL18*^87^; previously described EBV-induced neuronal genes *NRCAM* and *PPP1R17*^33^; and the metastasis-promoting gene *WASF3*^88,89^. Strikingly, we also observed elevated expression of *CD6* and *TNFRSF1A* in EBV^+^ and CIS atypical subsets relative to healthy control ABCs and conventional B cells (**Fig. 2D**, orange box). This finding was particularly intriguing, given that *CD6* and *TNFRSF1A* are MS susceptibility genes^90^. In particular, CD6 deficiency in mice has been shown to mitigate the severity of experimental autoimmune encephalomyelitis (EAE, a mouse model of MS) and, in T cells, impede immune infiltration of *in vitro* models of brain microvasculature and blunt IFNγ production/Th1 polarization^91^. We speculate that similar molecular functions of CD6 expressed by ABCs could contribute to the niche’s neuroinvasive and pro-inflammatory potential. Further comparison of *in vitro* EBV^+^ and CIS patient-derived ABCs is provided in a supplementary figure (**Fig. S3**).

Although certain genes expressed upon EBV infection were exclusive to CIS among disease datasets, GO analysis identified similar biological processes across *de novo* EBV infection and disease-state ABCs (**Fig. S4**). Relative to the resting phenotype, each disease ABC state was defined by significantly enriched terms related to viral transcription, gene regulation, and antiviral defense. As expected, terms related to immune cell signaling and activation were also uniformly observed. Notably, significant enrichment of genes associated with response to interferon gamma was observed in ABCs from MS, RA, SLE, and PSS, but not in either chronic infection context. TNF family cytokine production was enriched in ABCs from MS and SLE. Unexpectedly, expression of genes associated with neutrophil activation and degranulation were significantly enriched in ABCs from MS, SLE, PSS, and HIV.

This analysis defined disease-specific transcriptomic signatures of ABCs across autoimmune and chronic infectious diseases relative to EBV-induced expression. It is striking that a presumably idiosyncratic collection of genes upregulated in ABCs following EBV infection are also expressed in disease-associated ABCs but not those from healthy donors and that these transcriptomic similarities appears to be greatest in CIS. The apparent lineage-ectopic nature of this signature may be an artifact of unknown or uncharacterized functional roles for these genes in B cells. Alternatively, it is at least conceivable that their expression, not normally detected in B cells, may be associated with induced reprogramming to promote and maintain aspects of cellular plasticity (*ALDH1A1*, *POU5F1*, *MLLT3*, *NOTCH4*, and *HOXB4*)^92–99^ (**Fig. S5**). In this regard, it is arguably worth speculating that ABCs may be primed for even further aberrant phenotypic and functional singularity in response to complex stimuli including viral infection.

Several genes upregulated in EBV^+^ and CIS ABCs relative to healthy donors imply an enhanced capacity for chemotaxis and tissue invasion. For example, *WASF3* (which encodes an actin binding protein that regulates cytoskeletal morphology) promotes cellular metastasis and invasion in breast and prostate cancer contexts^88,89^. Likewise elevated *CXCR3* expression, which in T cells promotes migration from lymph nodes to inflammatory sites^100,101^, has been defined as a characteristic of neuroinvasive B cells in several contexts including MS^65,102,103^. Because CXCR3^+^ B cell frequency in MS is positively correlated with EBV viral load^65^ and the virus can induce B cell CXCR3 mRNA and protein expression^33,47^, EBV infection *per se* may promote ABC trafficking to the central nervous system. Likewise, the observed induction of *CD6* expression could conceivably facilitate the niche’s neuroinvasive propensity. In this regard, expression of neuronal genes involved in cell-cell interactions and axon guidance (e.g., *NRCAM*, *PPP1R17*, *PRX*) is further suggestive of a neurotropic capacity. Upregulation of *DLG4* within EBV^+^ and CIS ABCs is also noteworthy, as this gene encodes a neuronal signaling regulator that was identified within MS patient leukocytes and exhibited significantly reduced expression during IFNβ treatment^104^. ABCs even from healthy individuals basally express Src family kinases (*HCK*, *FGR*), which are essential to myeloid cell inflammatory responses^105^, but also inflammatory response inhibitors (e.g., *CD200R1*)^106^. EBV infection has previously been shown to induce *FGR* expression in an EBNA2-dependent fashion^78,79^. Upregulated expression of *IL18*, the pro-fibrotic wound healing mediators *AREG*^107^ and *PDGFD*^108,109^, and the inflammation-mediating inhibitory receptor *CD300A*^110^ within EBV^+^ and CIS phenotypes further indicate potential functions or participation in (neuro)inflammatory responses.

Given the expression profiles of ABCs in CIS patients and their similarities with EBV^+^ ABCs, we further investigated two CIS clusters (CIS_1a and CIS_5) from the original 42 disease-wide clusters that contained appreciable numbers of *CD19*^+^/*MS4A1*^+^/*TBX21*^+^ cells (**Fig. 3A**). CIS_1a and CIS_5 contained 1,371 cells from 30 of 32 samples (15 of 16 patients), which were further partitioned into four subclusters and stratified by timepoint and trial outcome. This population contained 1,298 cells from all patients eventually diagnosed with MS during the 18-month study period (13/13; 100 ± 69 cells per patient) and 73 cells from 2 of 3 long-term non-progressors (LTNP; 24 ± 31 cells per patient). Of the four identified subclusters, cells in CIS_1a (n = 395) and CIS_5 (n = 17) exhibited the greatest transcriptomic similarity to ABCs infected with EBV *in vitro* (**Fig. S3**). The CIS_1a subcluster accounted for 1.8% of all peripheral B cells in the full CIS cohort (n = 20,839). Within CIS_1a, 19 cells originated from 2 of 3 LTNP patients (6.3 ± 5.7 cells per patient) and 376 originated from 13 of 13 eventual MS diagnoses (15 ± 14.7 cells per patient). Although the sample size of CIS_5 was small (n = 17 cells; 8 cells from 5 patients at t_1_ and 9 cells from 6 patients at t_2_), it represented data from seven individuals, all of whom were ultimately diagnosed with RRMS (e.g., 7/13 MS outcomes, 0/3 LTNP outcomes) (**Fig. 3B-C**). In addition to quantifying ABC cluster composition by patient outcome, we identified a limited number of genes that were differentially expressed between MS activity and LTNP groups (**Fig. S6A-B**). Genes with significantly elevated expression in LTNP ABCs included *PEX11G*, *ATL1*, *MTRNR2L8*, *CHI3L2*, *SIRPA*, and *EZH2*. Of these, *MTRNR2L8* has previously been identified as a biomarker with potential predictive diagnostic value in MS, with reduced expression correlating to disease progression^111^. Genes with significantly downregulated expression in LTNP ABCs included *CD200R1*, *TNF*, *CCDC30*, *SYP*, *POU6F1*, and *FOXO4*.

**Figure 3.**
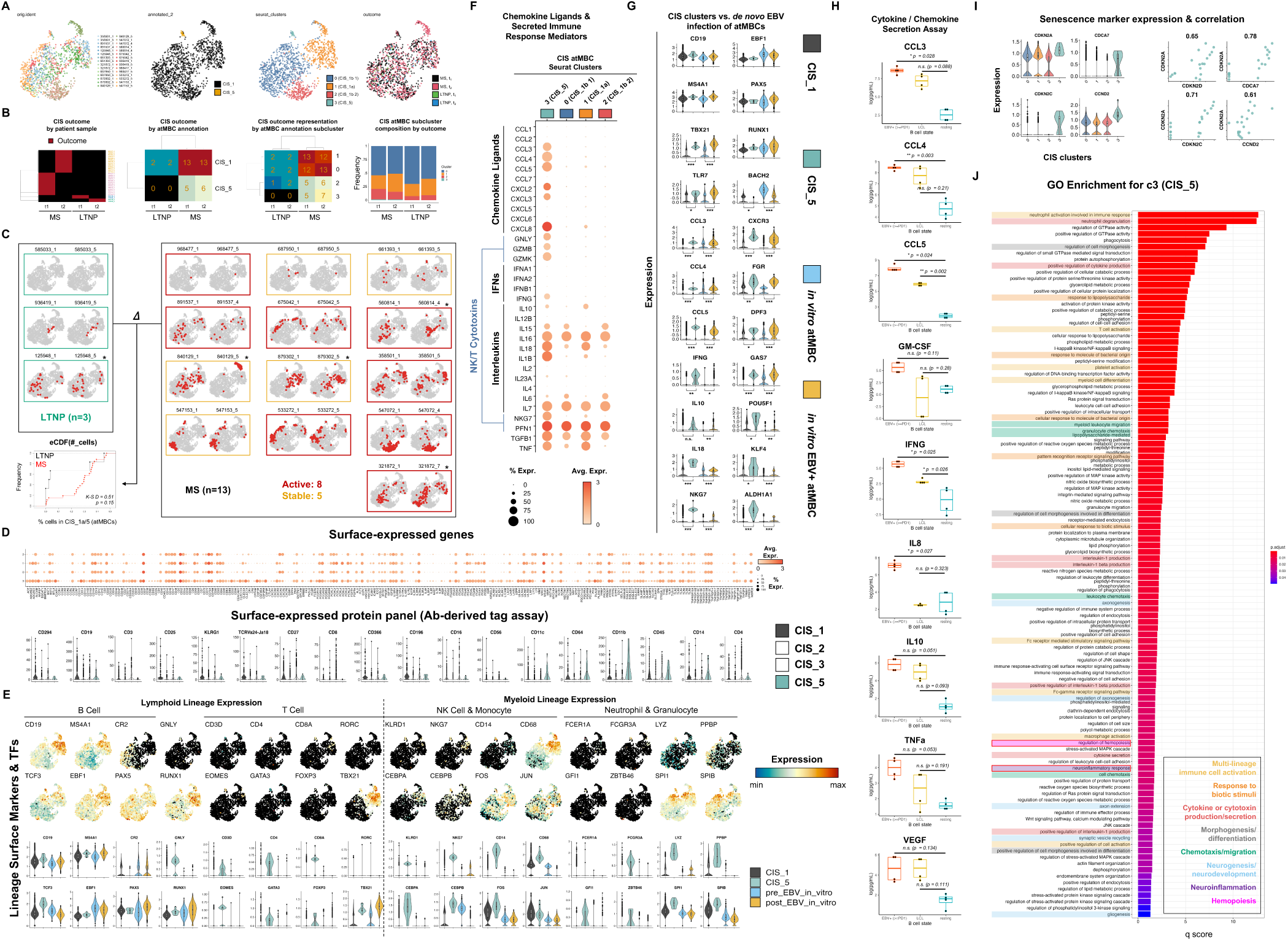
Detection of a rare pro-inflammatory ABC phenotype in CIS stratified by clinical outcome. **A)** UMAP visualization of CIS_1 and CIS_5 B cell clusters. Cells are plotted by original time-resolved patient sample (left panel), original (middle-left panel) and high-resolution (middle-right panel) clusters determined by unsupervised methods, and time-resolved clinical outcome (t_1_ and t_2_ from MS and LTNP stratification). **B)** Cluster composition by outcome-stratified patient samples. CIS sample IDs (left panel), number of CIS samples represented in original clusters (middle-left panel), number of CIS samples represented in high-resolution clusters (middle-right panel), and cell frequency in high-resolution clusters (right panel) for each timepoint and outcome. **C)** UMAP vizualization of all cell identities in CIS_1 and CIS_5 B cells by patient sample and timepoint. LTNP samples (green), MS active primary endpoint samples (red), and samples from CNS stable patients with post-trial MS activity (gold). Empirical cumulative distributions of cells in CIS_1 and CIS_5 by outcome and statistical comparison (n.s., Kolmogorov-Smirnov test, D-statistic = 0.51, p = 0.15). **D)** Non-canonical surface gene expression in ABCs from CIS cohort. Single-cell RNA-seq assay of surface-expressed genes in CIS_1 and CIS_5 B cells by high-resolution cluster (top panel) and CITE-seq (antibody-derived tag sequencing) of surface protein expression on CIS_1 and CIS_5 cells relative to two *TBX21^-^* B cell clusters (CIS_2, CIS_3) from the same cohort (bottom panel). **E)** Multi-immune lineage biomarker and transcription factor expression in CIS and *de novo* EBV-infected ABCs. In addition to *TBX21* expression, CIS_5 ABCs exhibit diverse immune lineage markers. **F)** Broad chemokine, cytokine, and cytotoxic scRNA-seq expression profile of CIS_5 ABCs. **G)** Significant shared scRNA-seq differential expression trends between CIS_5 and *de novo* EBV- infected ABCs indicative of immune activation, inflammatory response, and lineage plasticity. **H)** Secreted cytokine profiling of B cells in response to *in vitro* EBV infection. Data are presented for purified resting B cells (blue), proliferative B cells in early EBV infection (>=PD1, red), and latently infected cells (LCL, 5 weeks post-infection). Statistically significant differences in cytokine secretion were calculated using Welch’s two-tailed t-test from measurements (n = 4 biological replicates per condition). **I)** Correlated scRNA-seq expression of select genes associated with cellular senescence in CIS_5 ABCs. **J)** GO enrichment analysis for top differentially expressed markers of CIS_5 ABCs.

Although the difference in cumulative distributions of all ABCs (CIS_1a + CIS_5) from MS versus LTNP outcomes was not statistically significant (KS test p = 0.15, D statistic = 0.51), we sought to further investigate outcome representation in the CIS_5 phenotype. Because CIS_5 constituted a very limited sample size (n = 17 cells), we began by mapping all cells by patient ID (**Fig. S7A**) and cells from CIS_5 (**Fig. S7B**) back to the entire CIS B cell population (20,839 cells) to potentially identify a related cell neighborhood. Cells from CIS_5 remained tightly grouped as expected and comprised a subset of a larger cluster (c12, n = 81 cells) identified with unbiased methods. Accounting for the number of B cells derived from each sample, the six libraries from LTNP patients (3 patients x 2 timepoints) accounted for 4,259 (20.4%) of all B cells collected across the CIS cohort. In contrast, c12 contained only 4 B cells (4.9% of c12) from LTNP patient samples. Assuming unbiased sampling from whole blood, we calculated that the c12 phenotype represented 0.094% of peripheral B cells from LTNP patients versus 0.46% from patients ultimately diagnosed with MS, representing a 4.9-fold depletion in LTNPs (**Fig. S7C**). Of the 81 cells in c12, 77 came from 11 of 13 patients diagnosed with MS and 4 were from 2 of 3 LNTP. Poisson simulation determined that the expected number of cells in c12 derived from LTNP patients based on total patient B cell sample sizes and random chance was roughly 17 (λ= 16.6). Moreover, the cumulative Poisson probability of observing ≤ 4 cells (the empirical number from LTNP samples) was 2.5×10^-4^. This simulation was repeated to account strictly for *TBX21*^+^ cells within c12 (n = 63; 60 from 10/13 MS diagnoses, 3 from 1/3 LTNP) and produced similar results (# LTNP cells observed = 3; expected [λ] = 13; Λp[n β 3 cells] = 1×10^-3^) (**Fig. S7D**). Thus, with high probability, the c12 ABC phenotype (a superset of CIS_5) is significantly underrepresented within LTNP versus patients eventually diagnosed within MS.

Cells within CIS_5 exhibited an unusually broad set of surface-expressed genes at both the mRNA and protein levels (**Fig. 3D**). In the scRNA-seq assay, the most distinctive of these included *C5AR1*, *CD14*, *CD33*, *CD274* (PD-L1), *FCAR* (CD89), *HAVCR2* (CD366), *ITGAM* (CD11b), colony stimulating factor family members (*CSF1R*, *CSF2RA*, *CSF3R*), Fc gamma receptor family members (*FCGR1A* (CD64), *FCGR3A*, *FCGR3B*), killer cell lectin-like receptors (*KLRB1*, *KLRD1*, *KLRK1*), and classic T cell markers (*CD4, CD8A, CD8B*) in addition to ABC hallmarks (*CD19*^+^, *ITGAX*^+^, and *CR2*^lo^). This broad CIS_5 mRNA immunophenotype includes features (*CD19*^+^, *PTPRC*(CD45)^+^, *FCER2(*CD23*)^lo^*, *CR2*(CD21)^lo^, *ITGA4*(VLA4)^+^, *ITGAL*(LFA1)^+^, *TLR4*(CD284)^+^, CD5^int^) that are consistent with pro-inflammatory GM-CSF^+^ B cells described in MS^58^, which are closely related but not identical to innate response activator B cells in mice^112^. Results from the 18 marker CITE-seq assay identified protein-level co-expression of CD19, CD196 (CCR6), CD11b (ITGAM), CD11c (ITGAX), CD64 (FCGR1A), CD45 (PTPRC), and CD14 on CIS_5 cells, confirming the breadth of surface marker expression beyond a single classically defined immune lineage. Hematopoietic lineage marker and transcription factor profiling further supported this surprisingly diverse phenotype (**Figs. 3E, S6C-D**). It was noteworthy that, while EBV infection of ABCs *in vitro* led to elevated *TBX21* expression and modest expression of *NKG7* and *LYZ*, the multi-lineage immunophenotype of CIS_5 atypical cells was much more expansive than the previously described virus-induced phenotype. In this regard, cells within CIS_1a had greater transcriptomic similarity in the context of *de novo* EBV infection of ABCs.

The CIS_5 atypical state was further distinguished by elevated expression of a diverse array of genes encoding chemokine ligands, cytokines, and secreted factors with pro-inflammatory effects (**Fig. 3F-G**). These included C-C motif chemokine ligands (*CCL3* (MIP-1A), *CLL4* (MIP-1B), *CCL5* (RANTES)); C-X-C motif chemokine ligands (*CXCL2* (MIP-2A), *CXCL3* (MIP-2B), *CXCL8* (IL-8)); granzymes (GZMB, GZMK), *IFNG* (IFNγ); *IL10*, *IL18*, *IL1B*, *NKG7*, and *TNF* (TNFα). Several of these secreted factor genes (*CCL3*, *CCL4*, *CCL5*, *IL10*, *IL18*, *NKG7*) were also significantly upregulated at the mRNA level in the context of *de novo* EBV infection of ABCs (**Fig. 3G**). To further investigate potential contributions of EBV infection to cytokine production, we performed a 27-plex Luminex assay comparing secretion profiles of resting peripheral B cells, early EBV-infected cells, and EBV-immortalized lymphoblastoid cell lines (LCLs) (**Fig. 3H**). Levels of secreted CCL3, CCL4, CCL5, IFNG, and IL-8 were all significantly elevated in *de novo* EBV-infected proliferative B cells relative to resting cells. While not statistically significant, GM-CSF (p = 0.11), IL-10 (p = 0.051), and TNFα (p = 0.053) were also elevated upon EBV infection relative to resting B cells (Welch’s two-sided t-tests). Secretion of CCL5 and IFNG remained significantly elevated in LCLs.

We reasoned that the broad immune lineage plasticity and cytokine signatures exhibited by CIS_5 cells may be related to cellular senescence. Extensive expression of pro-inflammatory cytokines is consistent with the senescence-associated secretory phenotype (SASP), through which aged or damaged cells may promote immune cell recruitment in wound healing, regenerative, or developmental contexts^113^. Notably, senescent cells promote paracrine induction of cellular plasticity as well as “stemness” of the arrested cell via senescence reversal^114–117^. To this end, we evaluated the expression of hallmark senescence genes within CIS_5 cells relative to other CIS B cell subclusters (**Fig. 3I**). In addition to >85% of cells in G_0_/G_1_ based on cell cycle marker scoring (**Fig. S1**), CIS_5 cells exhibited upregulation of *CDKN2A (*p16^INK4A^), *CDCA7*, *CDKN2C (*p18^INK4C^), and *CCND2*. Further, expression of these four genes and *CDKN2D* (p19^INK4D^) were highly correlated. Collectively, these data support annotation of CIS_5 as a *CXCR3^+^*, pro-inflammatory ABC state, potentially associated with cellular senescence. Relative to other CIS B cell clusters, significantly enriched gene ontology terms for the top differentially expressed genes in CIS_5 cells reflect leukocyte chemotaxis, activation of multiple hematopoietic lineages, cytokine secretion, and, specifically, neuroinflammatory response (**Fig. 3J**).

It is important to emphasize that we did not detect EBV reads within B cell scRNA-seq data from the CIS patient cohort, which was also previously reported in a scRNA-seq study of patients with MS^49^. Given that EBV genomes and transcripts have been detected within MS brain lesions^59–64^ and correlate with neuroinvasive CXCR3^+^ B cells^50,65^, this absence may be due to one or more factors. These include transcriptome undersampling inherent to current scRNA-seq methods^118^; inefficient capture chemistry for the most abundant EBV transcripts (EBERs), which are not polyadenylated^119^; limited viral gene expression owing to restricted latency programs or failure to establish successful infection^33,120–122^; or a combination thereof. In this case, the transcriptomic profile exhibited by ABCs in CIS and in response to EBV infection *in vitro* may reflect epigenetically encoded responses of the cell niche that EBV, a prevalent stimulus, is capable of evoking. Likewise, although our prior work demonstrated that EBV can infect ABCs based on viral read detection^33^, we cannot definitively rule out the possibility that EBV-induced responses of other infected cells (e.g., inflammatory cytokine production) may indirectly activate ABCs.

In lieu of viral read detection, we used informatic methods to explore potential viral contribution to the observed CIS and EBV^+^ ABC phenotype. We used *cis*-regulatory prediction^123^ to identify genes associated with EBV nuclear antigen (EBNA) binding sites detected using ChIP-seq and cross-referenced these predictions against genes upregulated in CIS and EBV^+^ ABCs relative to resting cells (**Fig. S8A**). This yielded 82 EBNA-associated genes including *TBX21*, *CXCR3*, *FCRL2*, *FCRL5*, *FCGR2A*, *AREG*, *CCL5*, *CD69*, *CD84*, *CD300A*, *FGR*, *GPR174*, *HCK*, and *TBKBP1*. These 82 genes constituted modest enrichment of GO terms including defense response (GO:0006952, n = 17 genes, FDR = 0.0442), leukocyte activation (GO:0045321, n = 14 genes, FDR = 0.0442), regulation of cell migration (GO:0030334, n = 13 genes, FDR = 0.0468), and leukocyte degranulation (GO:0043299, n = 10 genes, FDR = 0.0468). Analysis of the *TBX21* locus in the lymphoblastoid cell line GM12878 revealed accessible chromatin with activating histone marks (H3K9ac, H3K4me3, H3K27ac), RNA pol II, and binding sites for EBNA-LP and EBNA-2 at the *TBX21* transcription start site (**Fig. S8B**). Collectively, informatic insights suggest that EBV-encoded transcriptional co-transactivators may mediate *TBX21*/T-bet expression and thereby affect ABC functions and responses. We also assayed expression of genes previously found to have high correlation to EBV lytic reactivation^124^ (**Fig. S9**). With few exceptions, host biomarkers of the EBV lytic phase were strongly and broadly expressed in CIS_5, the ABC subset with senescent hallmarks.

In summary, this study provides single-cell transcriptomic and protein-level evidence of ABCs with pathogenic features in patients with CIS. These cells, which constitute almost 2% of peripheral B cells, share many characteristics with ABCs in other disease contexts. Although we did not identify direct evidence of EBV infection within CIS ABCs, these cells were distinguished among diseases by transcriptomic similarities to ABCs in responses to EBV infection *in vitro*. A smaller subset of *CD19^+^*/*CD20^+^*/*TBX21^+^*/*ITGAX^+^* cells from patients with CIS (∼0.1% of B cells) that exhibited surprisingly broad immune lineage expression in addition to indicators of cellular senescence was absent or significantly underrepresented in long-term non-progressors. While definitive causality cannot be established in the absence of direct viral detection, the data presented here and prior work implicate EBV in a model wherein infection potentiates ABC migration and neuroinvasion by inducing expression of CXCR3^47,50,65^ and possibly other chemotactic receptors. The significant enrichment of neuronal genes including cell adhesion molecules in the EBV-associated ABC phenotype suggest an acquired neurotropic capacity that may facilitate CNS-localized viral antigen presentation and inflammatory responses that precipitate development of MS. Because ABCs differentiate into plasmablasts in response to innate and inflammatory stimuli^9^, it is further conceivable that EBV infection might promote low affinity antibody production by ABCs in the CNS. The likelihood of this pathogenic sequence of events would partially depend on the frequency of ABCs, which increases with age and to greater extent in genetic females^1^. In summary, this model is consistent with epidemiological aspects of the disease including age at time of EBV seroconversion and age- and sex-dependent accumulation of ABCs. Likewise, the clinical efficacy of anti-CD20 therapy^125^ and clinical exacerbations observed upon IFNγ treatment^126^ are each consistent with pathogenicity of CD20^+^ ABCs, which proliferate and differentiate in response to IFNγ. Notably, this model also aligns with empirical findings and prevailing hypotheses^55,127,128^ of EBV etiology in MS: molecular mimicry^56^, pro-inflammatory bystander damage^58^, and the potential for antibody production by autoreactive clones, the latter of which is a frequent fate of extrafollicular activated ABCs^12,13^.

It must be emphasized that this model and the data reported herein are not dispositive with respect to mechanistic involvement of EBV in CIS and do not preclude alternative causality. Specifically, there is a current lack of insight with respect to: 1) whether the observed age dependence of EBV seroconversion for risk in MS is directly related to the increase in ABC frequency over time; 2) whether EBV promotes or otherwise enhances the accumulation of ABCs *in vivo*; and 3) whether periodic EBV reactivation can result in chronic *de novo* infection of ABCs (or other subsets), consistent with autoimmune disease flares. Moreover, studies are needed to investigate the consequences of EBV infection in different genetic backgrounds with known autoimmune susceptibility variants. Thus, the nature of EBV infection in promoting the formation and activation of ABCs and the relevance of this host-virus relationship in MS and other autoimmune diseases warrants substantial focus in future work. Despite these unknowns, our findings underscore the value and importance of addressing questions of viral pathogenesis related to disease at cellular resolution with high-dimensional assays.

## METHODS

### Clinical study design (ITN STAyCIS Trial)

Patient PBMC samples used in this work were originally collected as part of the Immune Tolerance Network (ITN) STAyCIS Trial^70^. The clinical objective of the original STAyCIS Trial was to evaluate the safety and efficacy of Atorvastatin in CIS. A total of 81 patients with high risk of conversion to MS were enrolled upon initial CIS diagnosis in a two-arm clinical trial containing experimental (Atorvastatin, n = 49) and control (placebo, n = 32) treatment groups. The average age of the full cohort was 34 years, and the average time since CIS onset to screening was 65 days. All participants had no previous history of neurological disease and were seen within 210 days of the neurologic event. Patients samples were acquired at two timepoints following initial CIS diagnosis: t_1_ (baseline visit; at least 28 days after completion of corticosteroid treatment and within 210 days of CIS presentation) and t_2_ (at least 3 months post-t_1_; mean = 172 days post-t_1_, range = 84-357 days). Blood samples were collected and purified to PBMCs from each patient at each endpoint. We note that the criteria for MS diagnosis have been relaxed since the time of the STAyCIS study. Thus, by current standards some of the patients in this trial would have been diagnosed with MS at baseline.

### Patient samples for scRNA / CITE-seq

PBMCs from sixteen patients at each of two timepoints (n = 32) were selected for single-cell sequencing. Eleven of the selected patients were female (age = 32 ± 8.2 years) and five were male (age = 38 ± 6.4 years). Of these patients, eight met the defined primary endpoint for MS diagnosis within the 12 month treatment period and two participants met the endpoint criteria within 18 months from t_1_. Three additional participants subsequently met these criteria after the 18 month study period. The remaining three participants from the cohort analyzed herein were characterized as long-term non-progressors (LTNP) based on the absence of new T2 lesion, Gd-enhancing lesions, and no clinical exacerbations (3/16 selected cases).

### Single-cell library preparation

PBMCs from the 16 patients at each endpoint were incubated with a panel of 18 antibodies with unique conjugated sequencing tags (antibody-derived tags, ADTs) for surface protein expression profiling. The panel contained antibodies against the following surface markers: CD3, CD4, CD8, CD11b (ITGAM), CD11c (ITGAX), CD14, CD16 (FCGR3A), CD19, CD25 (IL2RA), CD27, CD45 (PTPRC), CD56 (NCAM1), CD64 (FCGR1A), CD196 (CCR6), CD294 (PTGDR2), CD366 (HAVCR2), KLRG1, and TCRVa24-JA18. Following ADT labeling, cells were prepared as single-cell RNA-seq (scRNA-seq) libraries using the 10x Genomics Chromium Controller and Single-Cell Gene Expression kit with v3.1 chemistry (5’ v1 single index). Cells were normalized to 1×10^3^ cells/μl, viability assessed, and titered to maximum of ∼10,000 cells per library. Cells are resuspended in master mix that contains reverse transcription reagents and combined with gel beads carrying the sequencing primers, barcodes, unique molecular identifier, and a poly-dT primer for RT. Full length cDNAs are cleaned and assayed to ensure lengths between 200-5000bp. Enzymatic fragmentation of the cDNA was used prior to adapter and sample index ligation; TruSeq read 2 primers are added via End Repair, A-tailing, Adaptor Ligation, and PCR. qPCR will be used to assess P5 and P7 adapter ligation, prior to size assessment of between 400-500bp. We generated 150bp paired end sequencing on an Illumina NovaSeq6000 (read 1 = 26 bp, read 2 = 91 bp, index read = 8 bp) at a target of 50,000 reads/cell for gene expression and 5,000 reads/cell for ADT libraries. Output base calls (.bcl format) from sequencing were then used to assemble single-cell RNA and ADT reads.

### Read assembly, QC, and alignment

We first demultiplexed raw FASTQ reads and aligned them to the human transcriptome before using Seurat^129^ to perform QC and analyze data by normalizing on a log scale after filtering for minimum gene and cell frequency cut-offs^130^. We identified and exclude possible multiplets^130^ and reduced noise by removing technical artifacts using regression methods^130^. Principal components (PCs) were calculated using the most variably expressed genes in our dataset^130^ and examined for cell dividing states^131^. Significant PCs were determined^131^ and carried forward for cell clustering and to enhance visualization^130^.

### Data processing and visualization

Upon completing the steps described above, we obtained ∼9,000 cells on average (8,949 ±1,559) from each of the 32 longitudinal CIS patient samples. CIS patient count matrices were prepared as a Seurat^132,133^ object containing 274,277 PBMCs after QC filtering to exclude cells with fewer than 200 unique feature RNAs. Data were log normalized and scaled prior to identification of the top 2000 variable features as per standard Seurat workflows. Normalized, scaled data were dimensionally reduced through principal component analysis (*RunPCA*). Cell neighborhood identification, UMAP projection, and unbiased clustering were performed (*FindNeighbors*, *RunUMAP*, *FindClusters*). Automated cell type annotation delineated 20,839 B cells (7.6% of PBMCs), which were considered for subsequent analysis. Adaptive low-rank approximation (ALRA) was used at this stage to generate an imputed gene expression matrix with preserved biological zeros (true absence of transcript expression) and correction of technical transcript dropout^134^. Imputed expression values are presented unless otherwise stated. Unbiased clustering () at various resolutions was repeated to identify and subset B cells based on *CD19*, *MS4A1*, and *TBX21* expression in downstream analyses. Cell cycle scoring and phase were also assigned based on expression of curated S-and G_2_/M-phase gene sets.

### Additional disease dataset provenance and curation

In addition to the newly generated CIS cohort data described above, we collected the following publicly available scRNA-seq datasets for autoimmune diseases and chronic infections: GSE133028 and GSE138266 (MS, CSF and peripheral B cells)^49,71^; GSE157278 (pSS, PBMC)^73^; GSE196150 (RA, synovial B cells)^74^; GSE135779 (SLE and healthy controls, PBMC)^72^; GSE149729 (Malaria, HIV, and healthy controls, peripheral B cells)^26^; and GSE189141 (*in vitro* EBV infection, peripheral B cells)^33^. Single cell count matrices from each of these studies were prepared as Seurat objects, from which B cell subsets were identified and extracted based on *CD19* and *MS4A1* (cells exhibiting ≥ 25^th^ percentile assay-wide expression of both genes). B cell scRNA-seq data from each of these datasets were processed and analyzed as described above for the CIS cohort data.

### Analysis, statistical methods, and simulation

Differentially expressed genes between cell phenotypes, disease diagnosis, unbiased clusters, and longitudinal outcome were identified using the *FindMarkers()* function in Seurat. For most comparisons, differentially expressed genes were returned if expressed in a minimum of 60% of cells in the group of interest with an average log_2_ fold change > 0.7. Differential expression data are included as supplementary data files, including calculated p-values with and without multiple hypothesis (Bonferroni) correction. We observed that some genes were identified as differentially expressed in select comparisons across diseases due to discrepancies in gene symbol annotation from publicly available datasets. Such genes were excluded from consideration and figure presentation. We note that this is a conservative approach that ensures the fidelity of presented data but may fail to identify additional differentially expressed genes across ABCs from different diseases. For certain analyses, expression of curated genes of interest previously identified from ABCs before and after EBV infection *in vitro* (using the same approach) were analyzed. For select genes of interest, statistical significance of differential gene expression across groups was calculated via Kolmogorov-Smirnov (KS) test. Poisson simulation was used to randomly sample cells from empirically observed cell frequencies per patient to evaluate outcome representation probabilities within specific phenotypes of interest. Cumulative Poisson probabilities were calculated for empirically observed cell frequency by phenotype in addition to expected frequencies (Poisson λ parameter).

### Gene ontology enrichment

Gene ontology (GO) biological process (BP) enrichment analysis was performed from differentially expressed gene sets using the *enrichGO* function from clusterProfiler^135^. Genes with log_2_ fold change values > 0.7 in a given comparison were considered for this analysis, using the whole genome as a background. Resulting terms were filtered to include those with Bonferroni-adjusted p < 0.05 and q < 0.1. Significantly enriched GO BP terms are presented as bar (barplot), dot (dotplot), and network (emmapplot) plots to represent significance and the number of genes per term set.

### Cytokine assays

27-plex Luminex cytokine/chemokine secretion profiling was performed from 48h supernatants of resting peripheral blood CD19+ B cells, early EBV^+^ B cells that had undergo at least one population doubling after infection (≥PD1), and EBV-immortalized LCLs (as in Price *et al.*^136^). Two technical replicates from each of two biological donors were prepared and analyzed for these experiments. Resting B cells were purified from PBMCs by negative isolation (BD iMag kit, BD Biosciences). Early EBV^+^ cells (≥PD1) were sorted by FACS at 6 days post-EBV infection of PBMCs via CD19 positivity and CFSE staining to track cell proliferation status. Sorted EBV^+^ cells were then re-cultured for two days, after which supernatants were harvested for analysis. LCLs were grown out from PBMC infections with limiting virus dilution and analyzed at five weeks post-transformation. Each of the three sample types were pure B cell populations. The cytokine panel detected the following secreted proteins: PDGFB, IL-1β, IL-1RA, IL-2, IL-4, IL-5, IL-6, IL-7, IL-8, IL-9, IL-10, IL-12, IL-13, IL-15, IL-17, CCL11 (Eotaxin), FGF2, G-CSF, GM-CSF, IFNγ, CXCL10 (IP-10), CCL2 (MCP-1), CCL3 (MIP-1α), CCL4 (MIP-1β), CCL5 (RANTES), TNFα, and VEGF. Statistically significant differences in cytokine secretion between resting B cells and ≥PD1 or LCLs were determined via KS test.

### ChIP-seq gene regulatory prediction

The Gene Regulatory Enrichment Analysis Tool (GREAT)^123^ was used to predict *cis*-linked genes to binding sites for EBNA-2, EBNA-3A, and EBNA-LP identified from ChIP-seq experiments in the GM12878 LCL^137,138^. Linked gene predictions were cross-referenced against genes that exhibited significant upregulated expression in both CIS and EBV+ ABCs relative to resting ABCs. Gene network analysis for EBNA-associated differentially expressed genes was performed using Cytoscape^139^, and EBNA as well as other epigenetic ChIP signals and bulk chromatin accessibility^140^ were visualized using IGV^141^.

## DATA AVAILABILITY

All clinical data associated with the STAyCIS Trial is publicly available on the TrialShare website. B cell scRNA + CITE-seq data from the CIS cohort reported herein will be made available to reviewers upon request and made public upon manuscript acceptance.

## Data Availability

All data produced in the present study are available upon reasonable request to the authors

## ACKNOWLEDGMENTS

The authors wish to thank Jay Tourigny for assistance with cytokine secretion profiling experiments reported herein and the Immune Tolerance Network (ITN) for thoughtful feedback in the development of this manuscript. We also thank Gillian Horn and Nico Reinoso in the Luftig laboratory for discussion and feedback.

## AUTHOR CONTRIBUTIONS

E.D.S., S.G.G., and M.A.L. conceived and designed the study and wrote the manuscript. E. Haukenfrers, V.J., K.A., & E. Hocke performed single-cell experiments and curated public datasets. E.D.S. and V.J. processed the data and extracted B cell populations. E.D.S. analyzed the data. L.A.C., K.M.H., and S.S.Z. reviewed and edited the manuscript. All authors have approved the final version of the manuscript.

---

**Figure S1.**
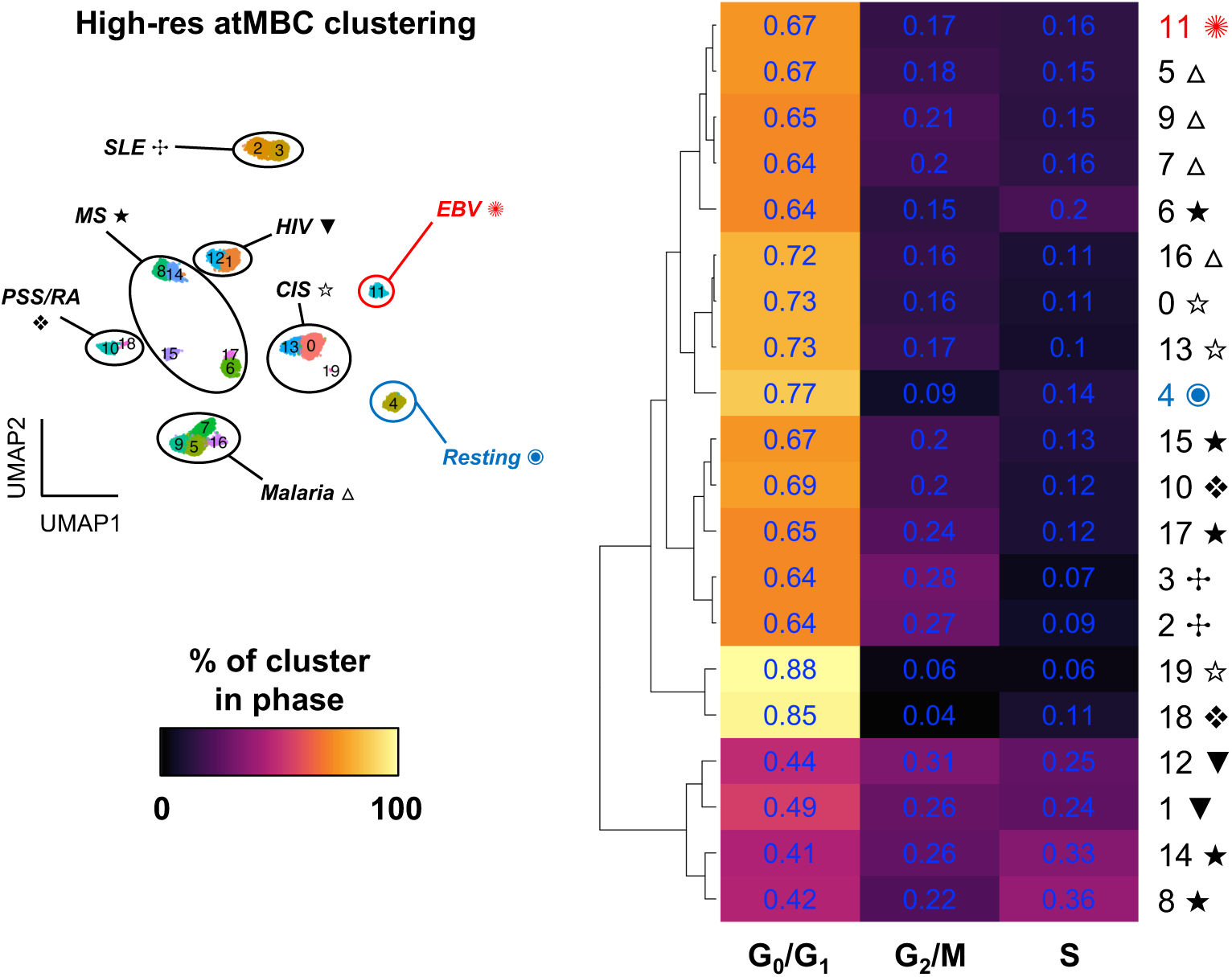
Cell cycle scoring of ABCs by disease.

**Figure S2.**
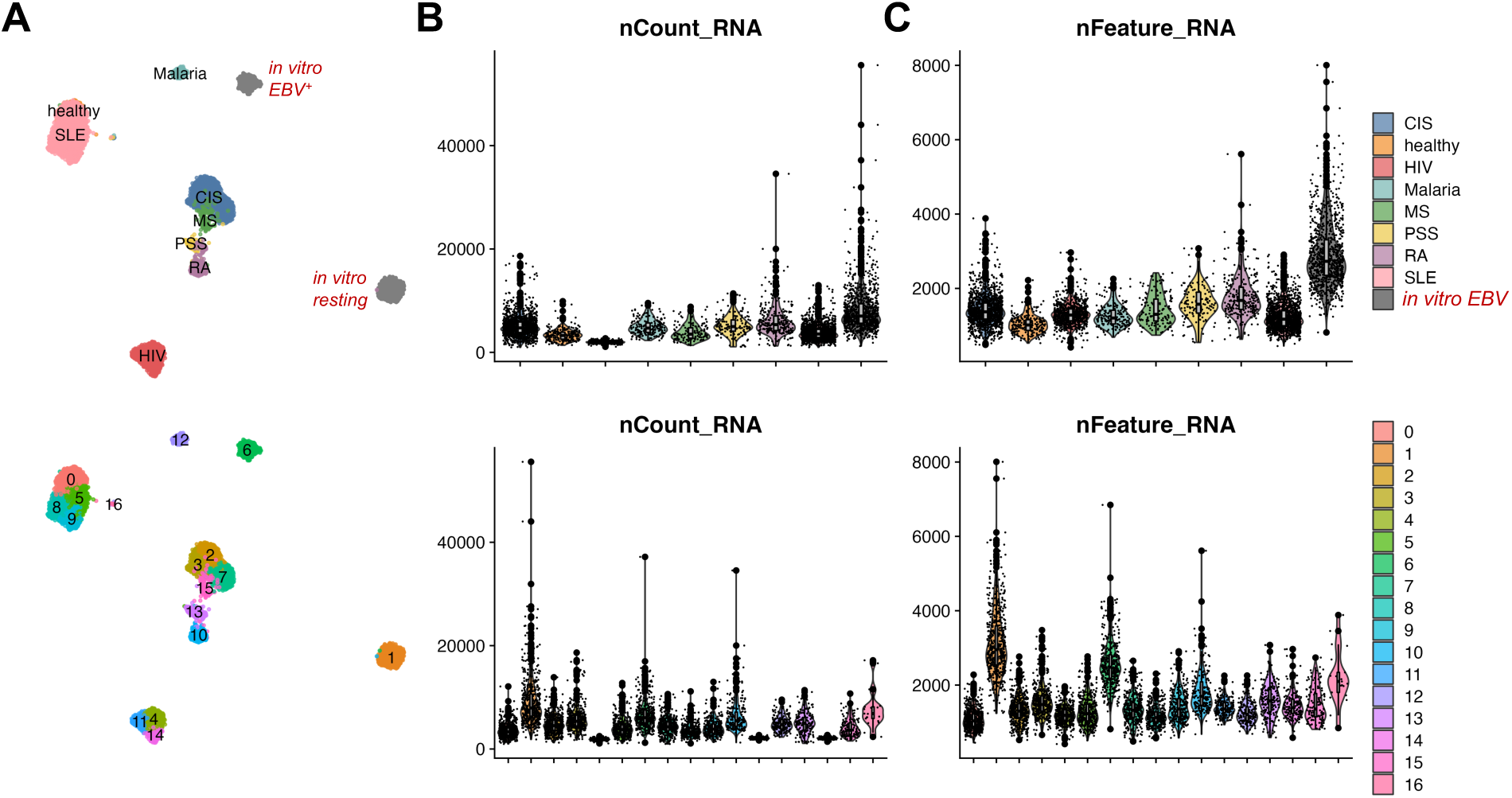
Global feature characterization of B cell clusters by disease dataset. **A)** UMAPs of select B cell phenotypes by disease. **B)** Distribution of total captured mRNA per cell by disease and cluster. **C)** Distribution of unique features (genes) per cell by disease and cluster.

**Figure S3.**
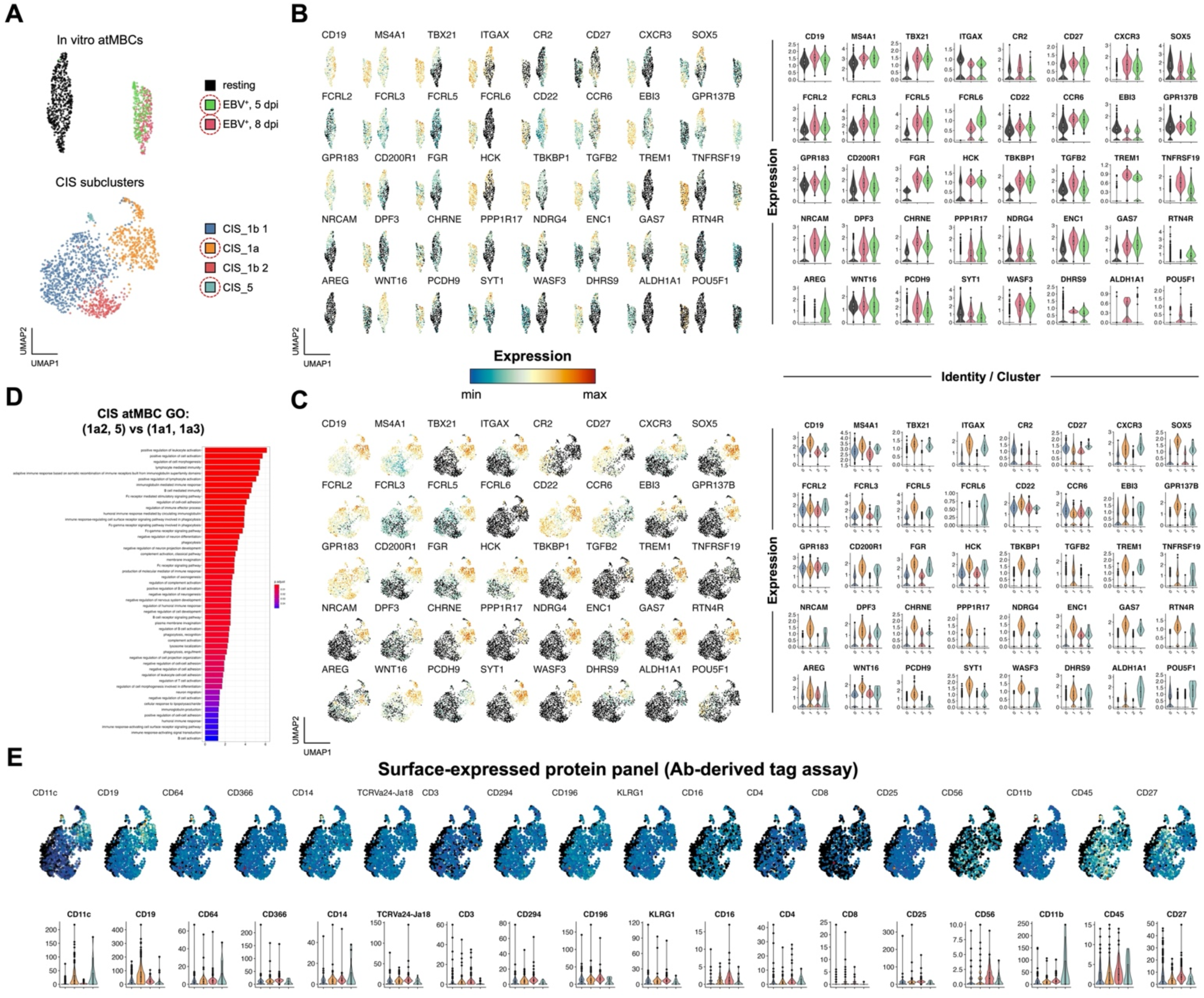
Detail of EBV-induced ABC signature in *de novo* EBV and CIS datasets. **A)** UMAPs of ABCs from time-resolved *de novo* EBV infection (top) and CIS cohort (bottom). **B)** UMAP and violin plot depiction of ABC gene expression changes in response to EBV infection. **C)** UMAP and violin plot depiction of EBV-induced genes shown in B within ABC clusters from CIS dataset. **D)** Gene ontology for CIS ABCs clusters (*TBX21*^+^; CIS_5, CIS_1a2) versus non-ABCs. **E)** UMAPs and violin plots of surface protein expression in high-resolution CIS clusters.

**Figure S4.**
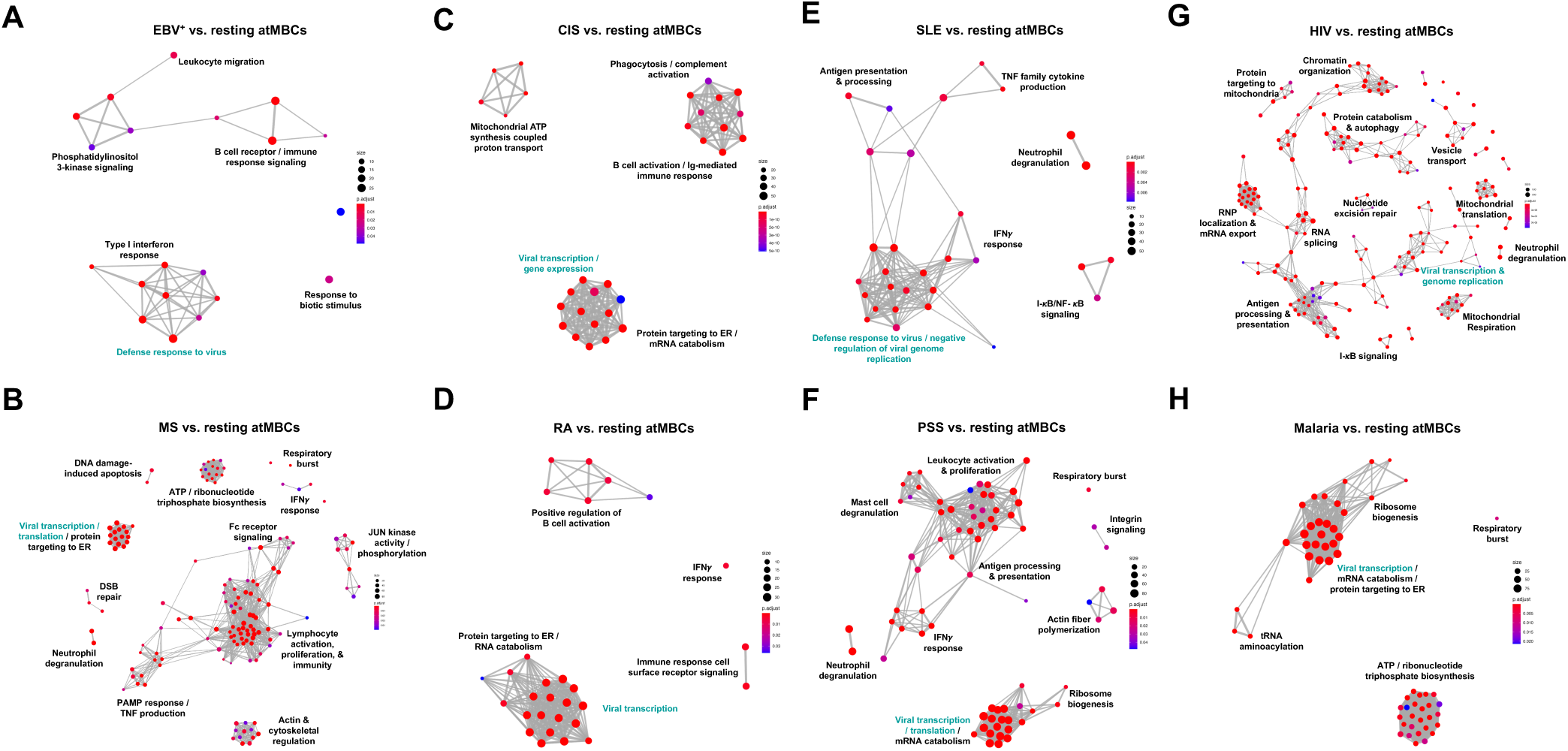
Gene network ontology for ABCs by disease. **A)** EBV^+^ versus resting ABCs. **B)** MS versus resting ABCs. **C)** CIS versus resting ABCs. **D)** RA versus resting ABCs. **E)** SLE versus resting ABCs. **F)** PSS versus resting ABCs. **G)** HIV versus resting ABCs. **H)** Malaria versus resting ABCs.

**Figure S5.**
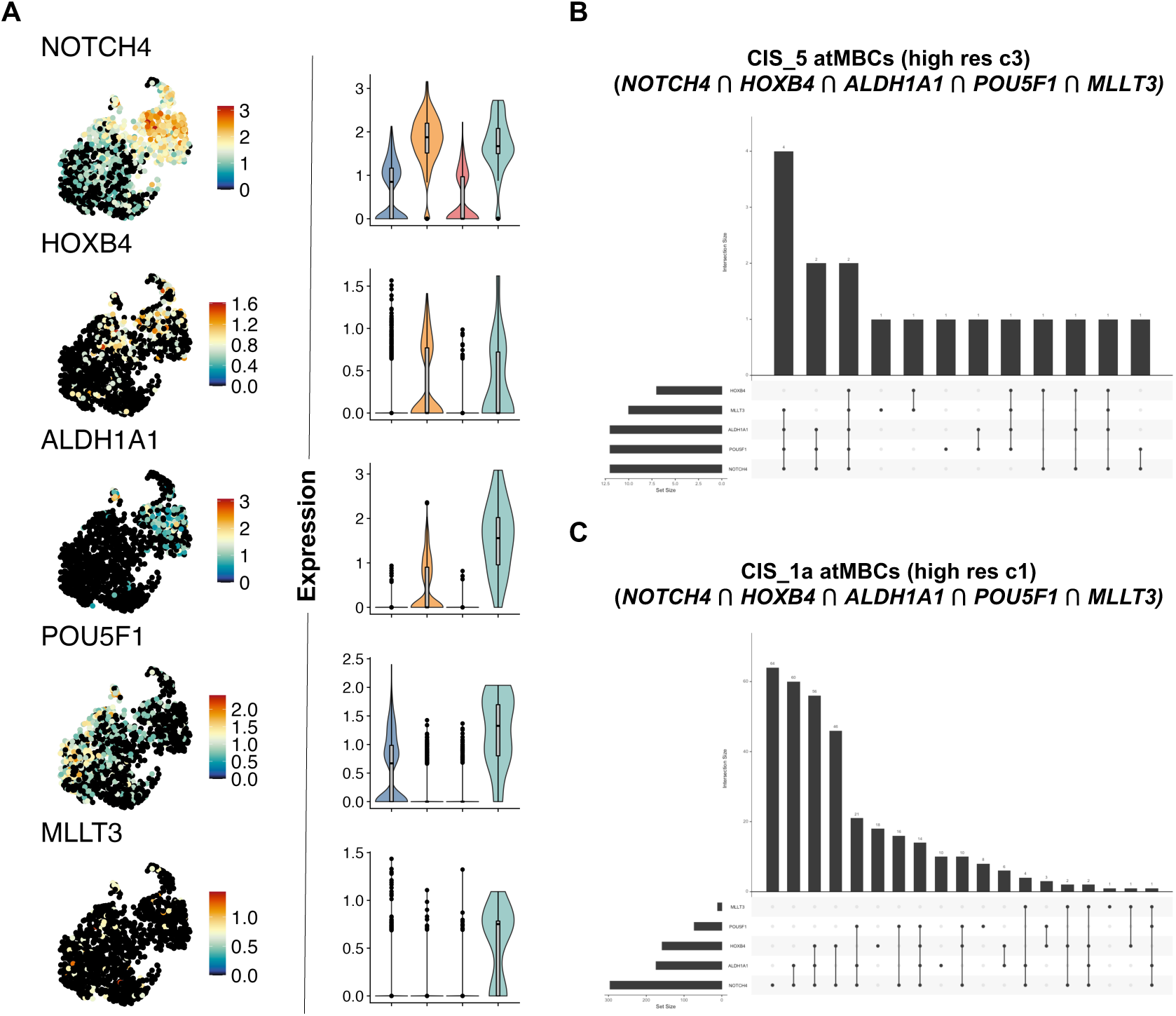
Co-expression of genes involved in maintenance or induction of cellular plasticity within CIS ABCs. **A)** UMAP and violin plots of gene expression by high-resolution CIS cluster (CIS_5 depicted in sage green in violin plots). **B)** Upset plot depicting co-expression in CIS_5 cells. **C)** Upset plot depicting co-expression in CIS_1a cells.

**Figure S6.**
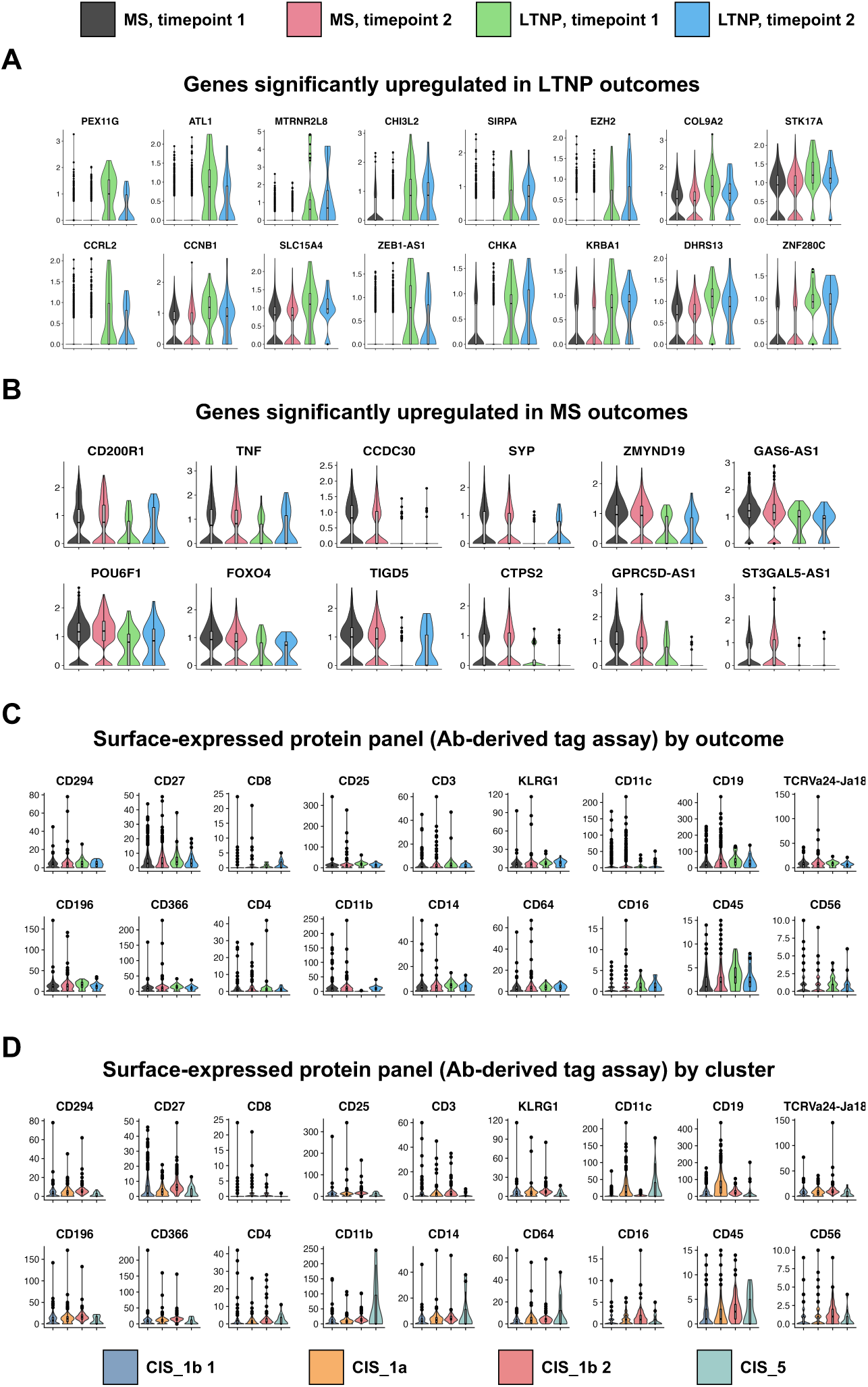
CIS B cell marker genes and surface protein expression by clinical outcome. **A)** Top genes enriched in B cells from LTNP samples versus MS samples. **B)** Top genes enriched in B cells from MS samples versus LTNP samples. **C)** Surface protein expression (CITE-seq assay) stratified by MS / LTNP outcome and sample timepoint. **D)** Surface protein expression stratified by high-resolution CIS ABC clusters.

**Figure S7.**
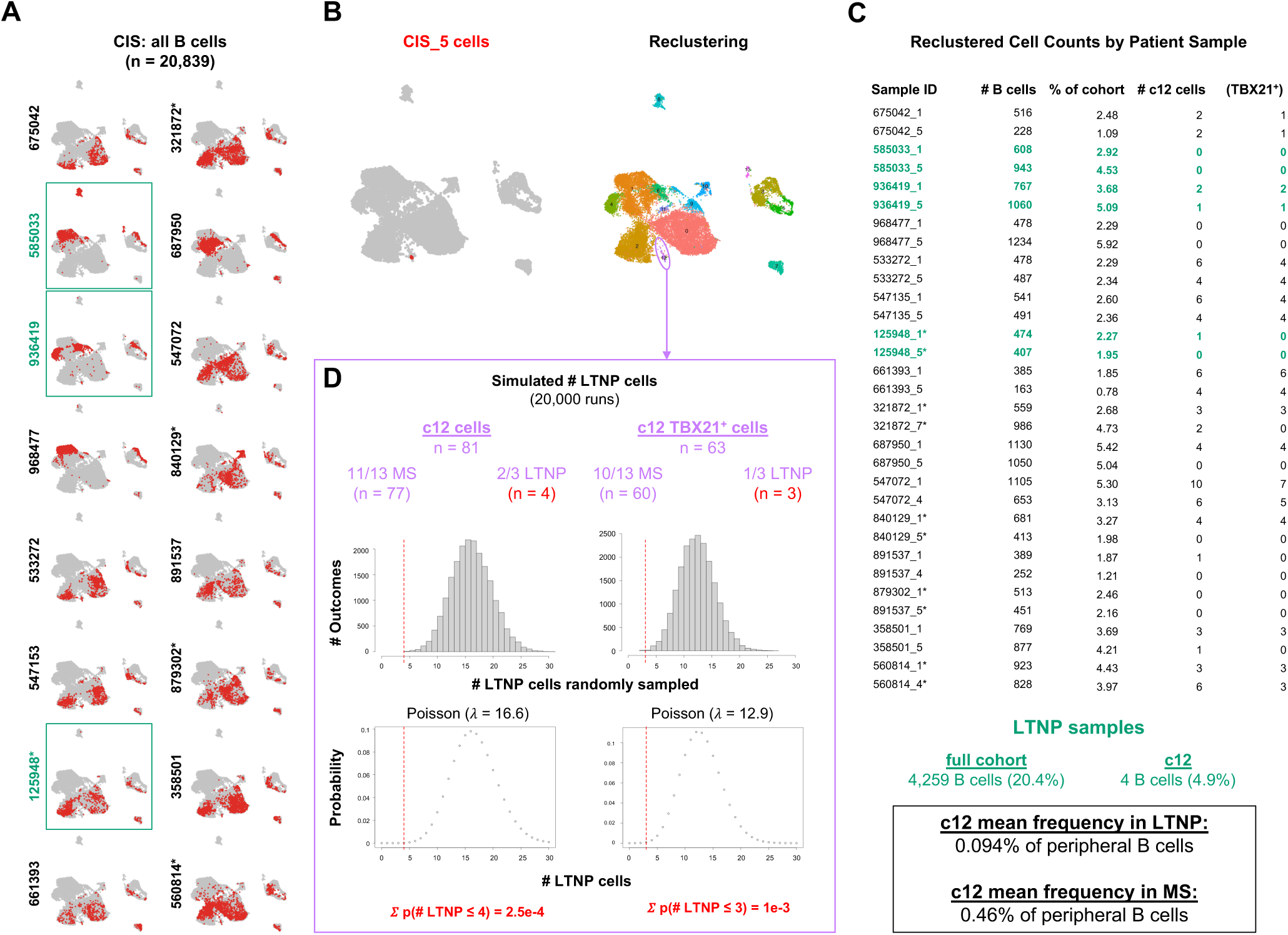
CIS_5 phenotype back-gating to all cohort B cells and associated phenotype frequency by clinical outcome. **A)** Patient-resolved B cell identities across CIS cohort samples. LTNP samples are depicted in green. **B)** Location of CIS_5 ABCs (red) within all CIS cohort B cells (left panel) and re-clustering to identify related phenotype (cluster 12, depicted in lavender; right panel). **C)** B cell representation by patient across full cohort and cluster 12. LTNP samples are depicted in green. Empirically observed frequencies of cells in cluster 12 are presented by clinical outcome (bottom panel). **D)** Poisson simulation comparing observed versus expected number of cluster 12 cells derived from LTNP samples based on total B cells per patient. For each outcome (MS, LTNP), the number of patients with cells present in cluster 12 as well as the number of cells per outcome are presented. Cumulative probability of observing the empirical number of cluster 12 cells (or fewer) from LTNP samples by random chance is depicted in red.

**Figure S8.**
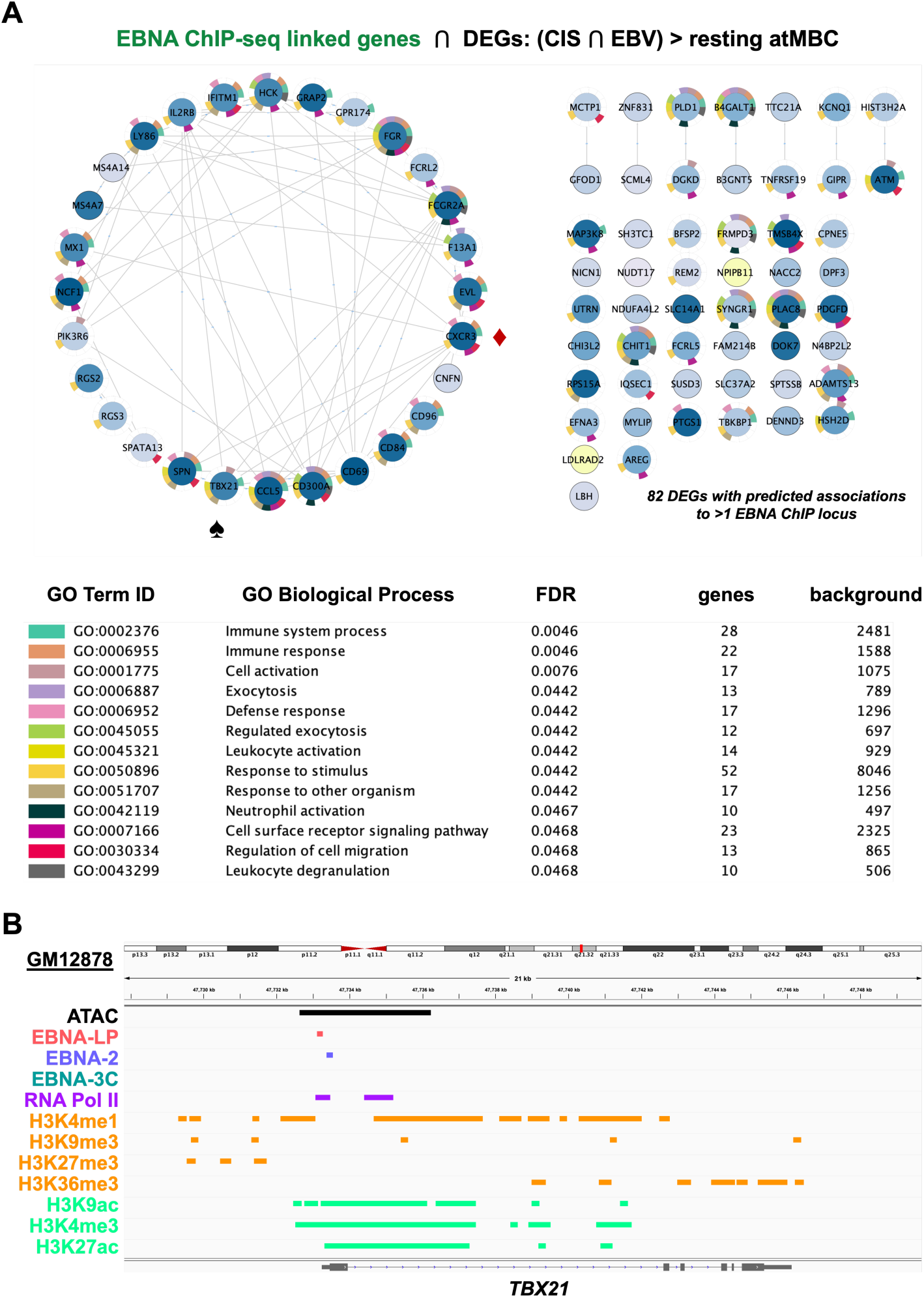
Genes expressed in ABCs from CIS and *de novo* EBV datasets with predicted *cis*-regulatory linkage to EBV nuclear antigen (EBNA) binding sites. **A)** Gene network representation and ontology of EBNA binding site-associated genes with elevated expression in CIS and *de novo* EBV-infected versus resting ABCs. These EBNA-linked differentially expressed genes include *TBX21* (T-bet) and *CXCR3* (denoted by spade and diamond icons, respectively). **B)** ATAC-seq and ChIP-seq data from the GM12878 cell line (EBV-transformed B cells) identifying open chromatin, EBNA binding sites, and epigenetic marks for active transcription at the *TBX21* transcription start site.

**Figure S9.**
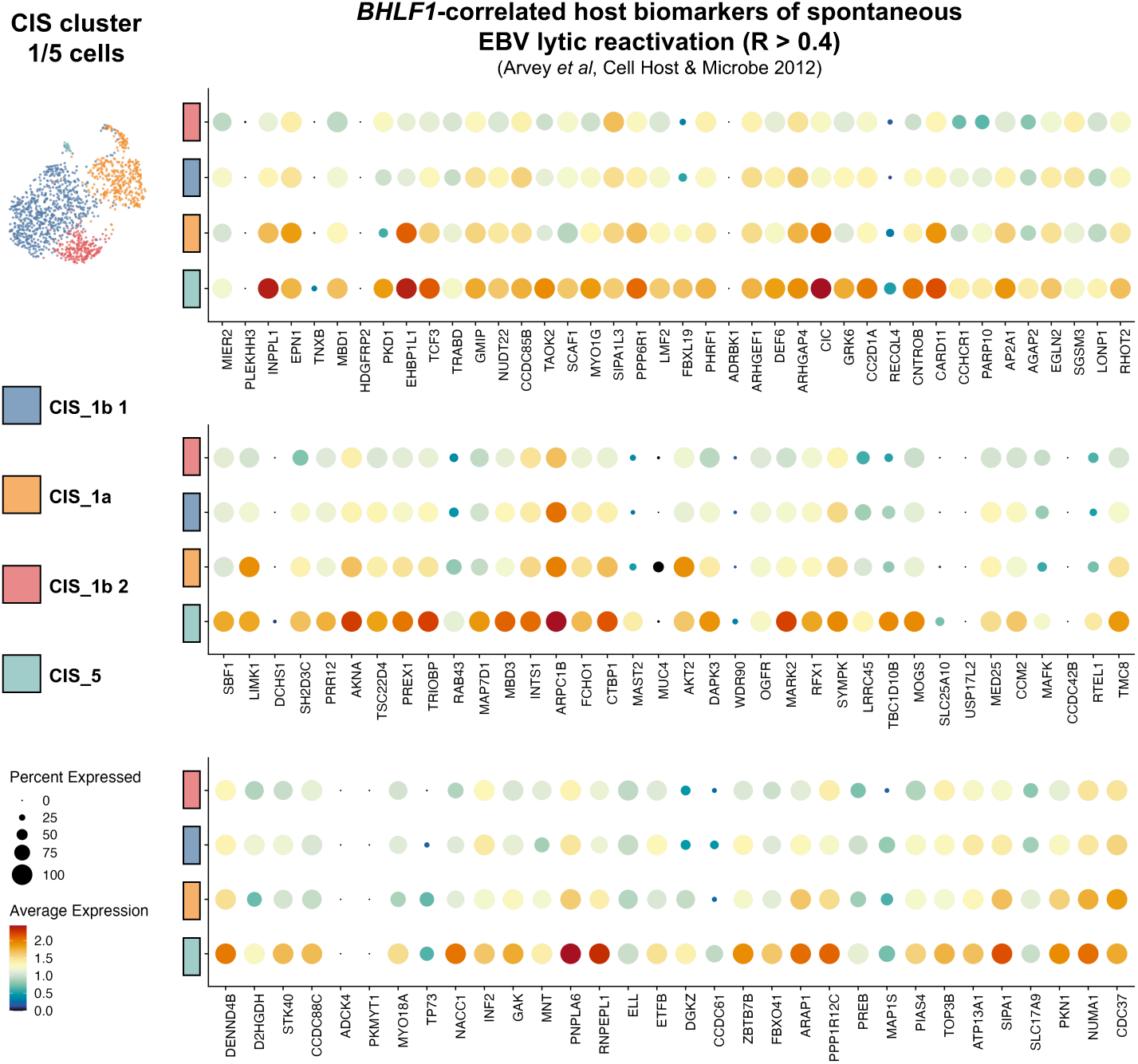
Expression of host cell biomarkers of EBV lytic reactivation from latency by CIS ABC cluster.

